# Are Dutch General Practitioners willing to prescribe mifepristone and misoprostol?: a mixed-methods study

**DOI:** 10.1101/2022.02.24.22270908

**Authors:** Julia E A P Schellekens, Claire S E Houtvast, Peter Leusink, Gunilla Kleiverda, Rebecca Gomperts

## Abstract

**Background:** The World Health Organization (WHO) indicates that General Practitioners (GPs) can effectively and safely provide mifepristone and misoprostol for medical termination of pregnancy (TOP). Dutch GPs are permitted to treat miscarriages with mifepristone and misoprostol, but in practice only guide spontaneous miscarriages. Current Dutch abortion law forbids GPs to prescribe these medications for medical TOP. Medical TOP is limited to the specialized settings of abortion clinics and hospitals. A shift to primary care is debated in the House of Representative, following the example of France and Ireland. It would improve reproductive health care and choices for women. Little is known about GPs’ willingness to provide medical TOP and miscarriage management.

**Aim:** This study aimed to gain insight into Dutch GPs’ willingness and anticipated obstacles to prescribing mifepristone and misoprostol for medical TOP and miscarriages.

**Design and Setting:** This is a mixed-method study among Dutch GPs.

**Method:** A questionnaire provided quantitative data that was analysed using descriptive methods. Thematic analyses were performed on qualitative data collected by in-depth interviews.

**Results:** The questionnaire was sent to 575 GPs, the response rate was 22.1%. Of the responders, 84.3% were willing to prescribe mifepristone and misoprostol and 58.3% were willing to provide both medical TOP and miscarriage management. 57.5% indicated a need for training. The main barriers influencing GPs’ willingness were lack of experience, knowledge, time and a restrictive abortion law.

**Conclusion:** Over 80% of the respondents were willing to prescribe mifepristone and misoprostol for medical TOP or miscarriages. Training, (online) education and a revision of the abortion law are recommended.

**How this fits in:** Medical TOP in the Netherlands can only be provided in abortion clinics and hospitals. GPs may prescribe these same medications for miscarriage management, but in practice only guide spontaneous miscarriages. To improve access to woman-centred care, it is important to allow GPs by law to provide medical TOP. Our study is the first to assess Dutch GPs’ willingness to provide mifepristone and misoprostol and aims to understand enablers and barriers that give insight into the feasibility of a shift in care. Our results illustrate the need to revise laws and to provide training and education in the similar procedure of medical TOP and miscarriage management.

## Introduction

Sexual and reproductive health is a basic human right and should be acknowledged by all. Every woman has the right to choose the number, timing, and spacing of her children in a free and responsible manner, without any discrimination, violence, or coercion. To realize these rights, access to legal, safe and comprehensive abortion care is essential (1, 2). General Practitioners (GPs) are at the centre of the Dutch healthcare system and function as gatekeepers to specialist care. Reproductive health, including contraceptive care, is part of their responsibilities. Historically, pregnancy-related care belonged to the domain of the GP but has shifted to specialised care. GPs can prescribe mifepristone and misoprostol for miscarriage management, but few actually do this. GPs are however not allowed to prescribe these same medications to terminate a viable pregnancy when women ask for medical termination of pregnancy (TOP). According to the Dutch abortion law, medical TOP can solely be provided in special clinics or hospitals. There are only 15 clinics that are all located in urban areas; two provinces do not have an abortion clinic at all. It is known that this is one of the many reasons that Dutch women experience obstacles to access abortion care (3–5).

Left-winged parties have submitted an amendment on the abortion law to allow GPs to prescribe these medications for medical TOP and thereby increase accessibility of abortion care and autonomy of women with an unwanted pregnancy. This amendment is discussed in February 2022.

The WHO’s safe abortion guidance indicates that GPs have the ability to effectively and safely provide mifepristone and misoprostol for medical TOP up to nine weeks and miscarriage management (6, 7). Dutch guidelines about miscarriage management indicate the same (8). GPs already prescribe mifepristone and misoprostol for medical TOP in several other countries, including France and Ireland, showing positive results (9–11). In these countries, both GPs and patients report to be satisfied and that it is effective and safe (12). Women report more control, anonymity and privacy, and it is less expensive (11, 13). The same was found in the United States, where primary care physicians were allowed to prescribe medical TOP after online consultation due to the COVID-19 pandemic (14).

Each year, about 30,000 women in the Netherlands terminate their pregnancy (15). The TOP rate is 9.1 per 1000 women living in the Netherlands aged 15 to 49, which is low compared to TOP rates worldwide and in countries with a similar healthcare system. This is thought to be the result of comprehensive sexual education and accessibility, availability and affordability of contraceptives, often provided by GPs (16). When contraceptives fail, they are regularly the first point of contact and two third of Dutch women visiting an abortion clinic are referred by their GP (15). It is hypothesized that many women would prefer to only visit their GP when seeking medical TOP or miscarriage management and not an abortion clinic or hospital (17).

It is clear that women benefit from the possibility to receive medical TOP and miscarriage management by GPs as it increases physical and mental autonomy (18, 19). A shift to primary care would eliminate many barriers they are currently experiencing when accessing abortion care. The crucial element to make a shift in care work is that GPs need to be willing to prescribe mifepristone and misoprostol for both medical TOP and miscarriages. Data on the overall willingness to provide this treatment and barriers perceived by Dutch GPs are lacking. This study aimed to gain more insight into the willingness of GPs in the Netherlands to provide mifepristone and misoprostol for medical TOP and miscarriages.

## Method

A mixed-methods study design was chosen, including both quantitative and qualitative methods of data collection and analysis. This study design allowed for complete analysis of the data, as both methods complement each other. For the quantitative part of the study, a 21-items multiple-choice questionnaire was conducted to gain insight into demographics, current practices and theoretical factors influencing willingness to provide medical TOP and miscarriage management. The questionnaire was based on a conceptual framework obtained from literature on abortion care and possible perceived barriers. For validation, the questions were sent to a pilot group and adapted based on their feedback. At the end of the questionnaire, all GPs were asked if they wanted to participate in a follow-up in-depth interview. When willing to participate, they could leave their contact details. The interviews were taken until data saturation was reached.

For the qualitative part of the research semi-structured interviews, all by phone, were conducted which allowed for in-depth topic discussion. The interviews were done by JS or CH, both performed five interviews. At the time of the interviews, they were doing an International Public Health master and Medicine master respectively; both had a personal interest in abortion care and strong pro-choice sentiment. A general interview guide with predetermined topics was created with room for personal stories. The questions were related to personal experience with medical TOP and miscarriage management and a possible shift of care. A pilot interview checked the quality of the interview guide. Interviews were recorded and transcribed. Investigator triangulation, performed by JS, CH and RG, reduced observer bias. Thematic analysis was performed to describe data in detail and identify patterns and emerging key themes.

### Participants sample

A systematic sampling strategy was used. Ideally, a GP from each municipality in the Netherlands would complete the questionnaire to create a diverse and representative sample. A list of all Dutch GPs with their contact details is not available due to privacy reasons. As this research was performed commissioned by a non-profit organisation, it was not financially viable to include a third party for data collection. Therefore at least one GP from each municipality, chosen randomly from the Dutch Chamber of Commerce KVK using zip codes, was approached; their data could be collected online. The questionnaire was sent via email and after two weeks, a reminder was sent. It was anticipated that not all GPs would fill in the questionnaire and therefore not all municipalities would be represented in the results.

The participants of the qualitative part were all included based on their willingness to participate. All GPs that left their contact details were sent a link to book a time slot for this interview; reminders were not sent.

### General Analysis

The Capability, Opportunity, Motivation and Behavioural model (COM-B model), developed by Michie, was used as a theoretical framework to assess willingness (20).

### Statistical analysis

Data screening was executed on a univariable level to exclude missing values and outliers. Internal consistency was measured. After data screening, all results were analysed using descriptive statistics. All statistical analyses were conducted with SPSS, version 24.0.

### Thematic analysis

Thematic analysis was performed on the qualitative data. All interviews we transcribed verbatim and coded in Atlas.ti. The six steps of thematic analysis as identified by Braun and Clarke were used (21). This COM-B model provided a thematic framework to classify the barriers and opportunities that were mentioned by the GPs into three themes: capability, opportunity and motivation. All codes were clustered into these three themes by two researchers (CH, JS), a third researcher (RG) could be asked for reassessment in case of disagreement. Barriers were defined as anything negatively influencing GPs’ willingness to provide mifepristone and misoprostol for both indications. Enablers were defined as anything positively influencing GPs’ willingness.

## Results

The first part of this mixed-methods study was a questionnaire. In total, 575 invitations were sent by e-mail to GPs with a participation request. The overall response rate to the questionnaire was 22.1% (127/575). 79.9% of the participants were female. 49.6% had been working as a GP for more than 15 years. 67.7% of the respondents worked in a general practice located in Utrecht or Noord-Holland. Table 1 describes the characteristics of all respondents of the questionnaires.

**Table 1:**
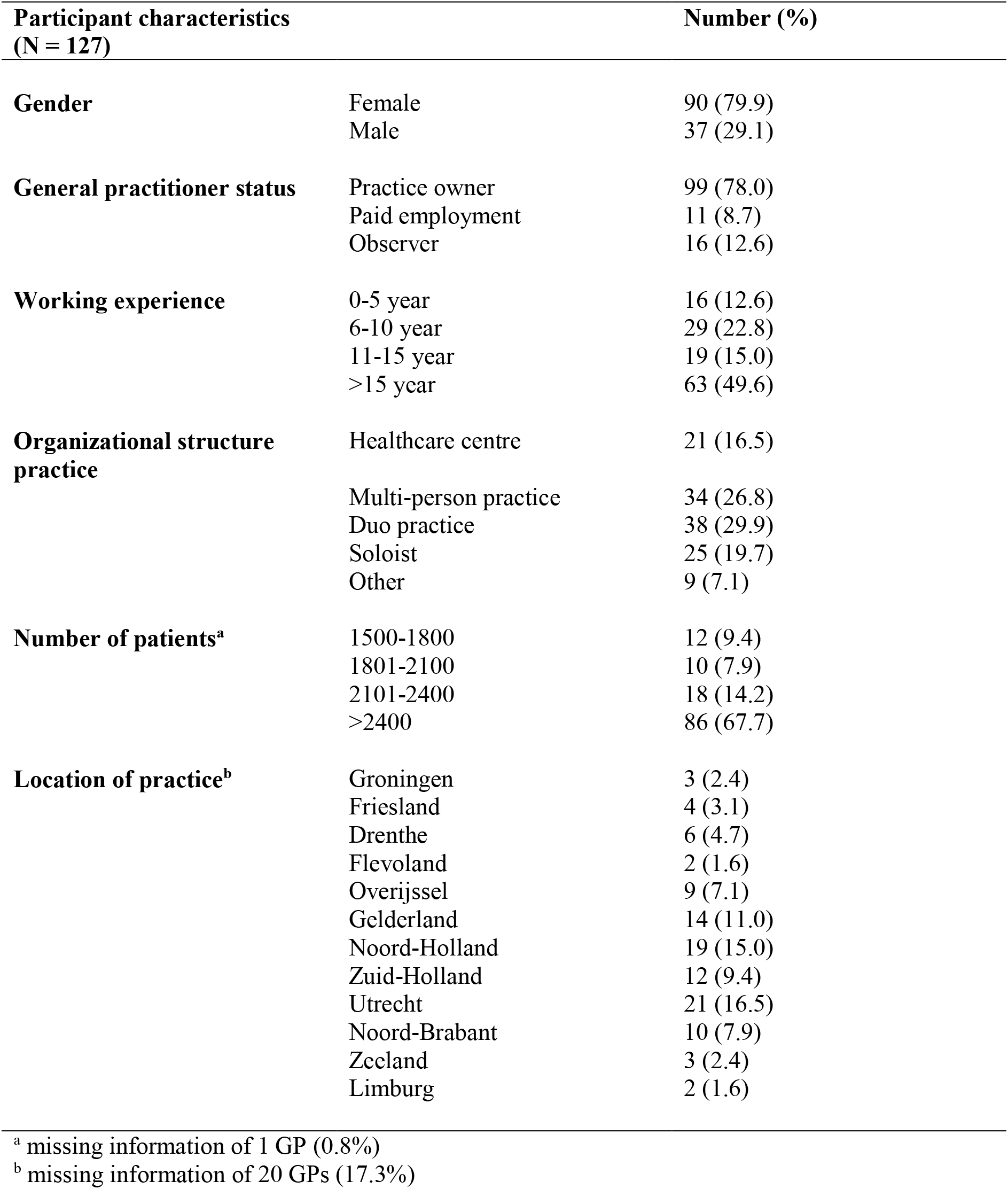
Participant characteristics of GPs that filled in the questionnaire.

The qualitative part of this mixed-methods study comprised an in-depth follow-up interview. Ten GPs, seven women and three men, from various Dutch provinces participated in these interviews; their characteristics are described in table 2. Table 3 describes the outcome of the questions concerning possible barriers and enables.

**Table 2:** Participant characteristics of the GPs of the interviews (N = 10)

| **Code of participant** | **Gender** | **Province** | **Organizational structure practice** |
| --- | --- | --- | --- |
| **R1** | F | Zuid-Holland | Multi-person practice |
| **R2** | F | Noord-Holland | Healthcare centre |
| **R3** | F | Friesland | Soloist |
| **R4** | F | Utrecht | Multi-person practice |
| **R5** | M | Noord-Brabant | Soloist |
| **R6** | F | Noord-Holland | Duo practice |
| **R7** | M | Gelderland | Duo practice |
| **R8** | M | Utrecht | Healthcare centre |
| **R9** | F | Noord-Holland | Soloist |
| **R10** | F | Utrecht | Duo practice |

**Table 3:** Outcome of the questionnaire.

| **Participant characteristics**<br>**(N = 127)** | **Number (%)** |
| --- | --- |
| **Miscarriage assistance request** |  |
| 0-2 times | 70 (55.1) |
| 3-5 times | 45 (33.4) |
| 6-10 times | 8 (6.3) |
| >10 times | 1 (0.8) |
| Not sure | 3 (2.4) |
| **Action method in case of a miscarriage** |  |
| Self-guidance and referral | 48 (37.8) |
| Preference to guide and treat self | 31 (24.4) |
| Direct referral to gynaecologist | 48 (37.8) |
| **Number of unwanted pregnancies annually** |  |
| 0-2 times | 59 (46.5) |
| 3-5 times | 54 (42.5) |
| 6-10 times | 13 (10.2) |
| >10 times | 0 (0) |
| Not sure | 1 (0.8) |
| **Action method in case of an unwanted pregnancy<sup>a</sup>** |  |
| Referral to abortion clinic, if no doubts | 70 (55.1) |
| Referral to abortion clinic, after discussion | 45 (35.4) |
| Against TOP, but will refer | 6 (4.7) |
| Against TOP, will not refer | 5 (3.9) |
| **Feeling qualified to provide medical TOP** |  |
| Yes, for a miscarriage | 6 (4.7) |
| Yes, for medical TOP | 4 (3.1) |
| Yes, for both miscarriage and medical TOP | 20 (15.7) |
| No, for neither miscarriage nor medical TOP | 97 (76.4) |
| **Willing to provide medical TOP after training<sup>b</sup>** |  |
| For miscarriage | 33 (26.0) |
| For miscarriage and medical TOP | 74 (58.3) |
| **Access to ultrasound** |  |
| Own practice | 9 (7.1) |
| Obstetrician's practice | 32 (25.2) |
| Other primary care diagnostics | 40 (31.5) |
| Referral to gynaecologist or abortion clinic | 46 (36.2) |
| **Need for additional training** |  |
| Yes, for miscarriages | 20 (15.7) |
| Yes, for medical TOP | 2 (1.6) |
| Yes, both for miscarriages and medical TOP | 73 (57.5) |
| No, neither for miscarriages, nor medical TOP | 32 (25.2) |
| **Barriers to provide medical TOP** |  |
| No barriers | 12 (9.4) |
| Extra administrative work | 12 (9.4) |
| Lack of experience | 75 (59.1) |
| Lack of time | 35 (27.6) |
| Lack of knowledge | 55 (52.0) |
| No access to ultrasound | 44 (34.6) |
| Objections from colleagues | 10 (7.9) |
| Lack of funding | 16 (12.6) |
| Personal conviction | 35 (27.6) |
| Public opinion (stigma) | 2 (1.6) |
| Other | 24 (18.9) |

<sup>a</sup> missing information of 1 GP (0.8%)
<sup>b</sup> missing information of 20 GPs (15.7%)

Figure 1 represents the participation rate in the different steps of this mixed-methods study.

**Figure 1:**
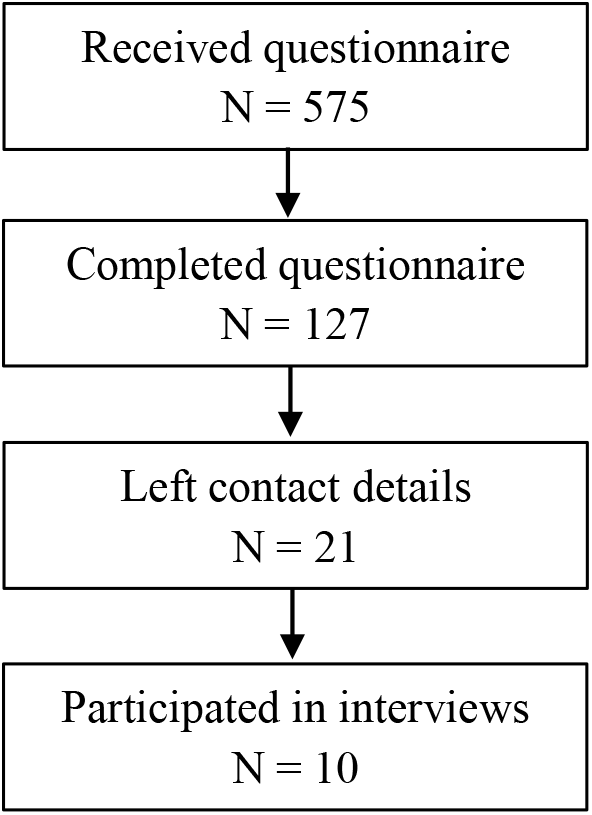
Flowchart on the participation rate of the quantitative and qualitative part.

### Capability

Lack of experience was mentioned by 75 (59.1%) GPs as a barrier to providing medical TOP and miscarriage management. Most of the respondents (76.0%) reported that at this moment they do not feel qualified to provide mifepristone and misoprostol for medical TOP nor miscarriages. Only 15.7% already felt capable enough to prescribe the medication for both medical TOP and miscarriages.

Lack of knowledge was considered a barrier by 52.0% of the respondents. 57.5% of the respondents indicated a need for training for both medical TOP and miscarriages. Only 25.2% of respondents did not wish to have any training. The interviews confirmed this lack of knowledge and information.

> *“I think it would be useful to receive proper education with people who think alike. So everyone willing to provide medical TOP should have the possibility to participate in class and get the information about all the ins and outs. You see, most things you already know, but it would never hurt to have someone with experience tell you what to expect*.*” (R5)*

> *“For example, for PrEP it is well organized. There are guidelines and a small summary chart available. That is how I would like to have it for medical TOP as well. I would prefer a plan that explains it step-by-step. I do not need to know all the details, but I do want to be able to look them up*.*” (R3)*

107 (84.3%) GPs indicated that they were willing to provide mifepristone and misoprostol after training; 26.0% would only prescribe the medication for miscarriages while 58.3% would be willing to prescribe mifepristone and misoprostol for medical TOP.

### Opportunity

Nearly half (55.1%) of the GPs reported seeing less than two miscarriages each year. 24.4% guide and treat miscarriages themselves while 37.8% immediately referred to a gynaecologist. Each year, unwanted pregnancies were seen less than two times by 46.5% and three to five times by 42.5% of the GPs. 58.3% of GPs were willing to provide mifepristone and misoprostol for medical TOP.

34.6% mentioned lack of access to ultrasound as a possible barrier. Although not necessary, 7.1% reported having access to ultrasound in their own practice, 25.2% had access to ultrasound diagnostics via a midwifery practice, 32.5% via other primary care facilities and 36.5% via referral to the gynaecologist or abortion clinic.

> *“For me, one of the main barriers is the fact that I do not have ultrasound equipment in our practice. I do have midwives that work with us, but they also do not have direct access to ultrasound equipment. If this was the case, then it would be much easier. Then I would probably arrange a direct link with one of the midwives*.*” (R4)*

A legal barrier mentioned several times was that it is in the Netherlands forbidden by law to provide medical TOP at the GP office. Several GPs suggested that the government should formulate clear guidelines on medical TOP by GPs and what tasks are included, to prevent misunderstandings or criminal offences.

> *“I do understand that GPs are not very enthusiastic about this idea. Nobody wants to end up at the disciplinary tribunal or have to undergo a juridical procedure. You should know for sure what does and what does not belong to your field of expertise*.*” (R2)*

### Motivation

Every interview participant believed that medical TOP and miscarriage management by GPs would be beneficial for women and increase access to care.

> *“I think it can be a nice opportunity to help women in difficult situations. Because after all, the GP is a place you can always go to in case of a care request*.*” (R10)*

Important motivational barriers were lack of time (27.6%) and lack of funding (12.6%).

> *“It is not the only task that has been added to our responsibilities. There are many tasks added and none are subtracted. We are supposed to provide more care for the same number of patients and if you look at our financing structure, we receive relatively little for the extra services that we provide because we still have subscription rates*.*” (R1)*

Stigma and fear of judgment about TOP were mentioned as motivational barriers by only 1.6% of the participants. However, for more than one quarter of the respondents (27.6%), personal conviction was a barrier to providing medical TOP.

Twelve respondents (9.4%) did not expect to experience any barriers to providing mifepristone and misoprostol.

## Discussion

This study aimed to explore the willingness of Dutch GPs to provide mifepristone and misoprostol for medical TOP and miscarriages by obtaining more insight into enablers and barriers that influence their willingness. The recent debates in the Dutch House of Representatives on the amendment to allow GPs to prescribe mifepristone and misoprostol for medical TOP makes this research valuable. During the interviews, it was known that some political parties were thinking about this amendment but they had not submitted it yet. The recent debates show the uncertainties on the perceived opportunities and barriers. The findings of this mixed-methods study can and should guide the policymakers once an amendment has been passed, and guide a revision on the guidelines on miscarriages by the Dutch College of General Practitioners.

This study showed that 84.3% of GPs were willing to prescribe mifepristone and misoprostol, and two-third of them are willing to provide medical TOP. Lack of experience, knowledge and time and a restrictive abortion law were the main barriers; (online) training, education and a revision of the abortion law can address those barriers.

We were able to include a diverse and heterogenic sample in the interviews. The mixed-methods design has proven to be a strength, as the qualitative data allowed us to gain deeper insight into what was highlighted in the quantitative data and consequently to further explore ideas and beliefs of GPs on this topic. Our interview sample consisted of ten participants and data saturation was reached after eight interviews. The quality of the interview guide was checked by executing a pilot interview. All interviews were performed following an interview guide, within one month. Thereafter all interviews were coded and investigator triangulation was used to reduce observer bias and improve inter-judge reliability.

This study has several limitations to consider. Firstly, due to a small sample size and low response rate, the generalizability of our quantitative results could be limited, which is a common characteristic of web-based surveys (22). In the Netherlands, there are 12.766 active GPs. In our sample, female GPs are slightly overrepresented, with 80.0%, compared to the 60.0% in the overall GP population (23). Secondly, TOP can be a controversial and sensitive topic and results should therefore be interpreted with caution due to possible a social desirability response bias. Thirdly, respondents for the interviews were sought based on their willingness to participate, causing a lack of randomization. None of the interviewed GPs had a negative attitude towards abortion care. It is probable that the interview answers do not represent the opinion of GPs who are sceptical about this topic. Lastly, the characteristics of the researchers could have influenced the interpretation of this research as they might be unconsciously biased when interpreting the respondents’ answers. To reduce potential bias a second researcher verified all results.

Our findings correspond to existing literature that reports lack of accurate knowledge and skills as a barrier to provide medical TOP and miscarriage management (24, 25). Therefore, appropriate training and online education tools for GPs to improve their affinity with the topic are necessary (26). For this reason, it should be incorporated into the medical curriculum (27). Research suggests that early exposure and education about abortion care will lead to a higher acceptance rate of medical TOP provision as being part of the range of responsibilities (28).

Respondents considered a lack of time as one of the main motivational barriers influencing willingness to provide medical TOP and miscarriage management. To overcome this barrier provision of training and education tools would be beneficial (29). GPs will develop more affinity with medical TOP and recognize that it does not need to be extra time consuming (30), especially since GPs indicated they are already involved in counselling and providing after care to women with an unwanted pregnancy or miscarriage.

Previous studies found that fear of stigma, judgement and negative reactions, influence physicians’ willingness to provide medical TOP (31), especially in conservative regions (32). However, our study found only a few participants mentioning stigma or fear of judgement as a barrier possibly due to the liberated climate in the country.

Lack of access to ultrasound diagnostics was another concern of Dutch GPs. However, it has been proven that an ultrasound is not required as a combination of a positive pregnancy test and the last menstrual period are efficient, safe and accurate to assess gestational age (33, 34). Furthermore, a direct referral network to a midwifery practice or gynaecology department that has access to ultrasound diagnostics already exists in case of uncertain gestational age or suspected ectopic pregnancy.

To our knowledge, this study was the first of its kind in the Netherlands to assess GPs’ willingness to provide mifepristone and misoprostol for medical TOP and miscarriages. The findings show that the majority of responding GPs are interested in providing this care. Policymakers have to address possible obstacles such as lack of experience, knowledge, time and a restrictive abortion law. Medical TOP and miscarriage management should become a part of the curriculum for GPs in training and online and onsite education during their careers.

## Supporting information

Decision of the Research Ethics Review Committee VU Amsterdam

## Data Availability

All data produced in the present study are available upon reasonable request to the authors.

## Ethical approval

The Research Ethics Review Committee of the Vrije Universiteit Amsterdam waived ethical approval for this work.

## Acknowledgements

We want to thank all GPs that took the time to complete the questionnaire or participate in the interviews.

We have chosen to use the terms ‘woman’ and ‘women’ in this systematic review, to ensure legibility. Using these terms, we do not mean to exclude people that can become pregnant but do not identify or feel comfortable with the word ‘woman’ nor women who have been unable to conceive.

## References

1. (WHO) WHO. Sexual and reproductive health [cited 2021 February 28]. Available from: https://www.euro.who.int/en/health-topics/Life-stages/sexual-and-reproductive-health/sexual-and-reproductive-health

2. (WHO) WHO. Preventing unsafe abortion. 2020.

3. Holten L, de Goeij E, Kleiverda G. Permeability of abortion care in the Netherlands: a qualitative analysis of women’s experiences, health professional perspectives, and the internet resource of Women on Web. Sex Reprod Health Matters. 2021;29(1):1917042.

4. De Graaf H. Seks onder je 25e: Seksuele gezondheid van jongeren in Nederland anno 2012 [Sex under the age of 25: Sexual health of youth in the Netherlands in 2012]: Eburon Uitgeverij BV; 2012.

5. Kleiverda G, Gomperts, R., Schellekens. M., & Leusink. P.. Abortushulp kent te veel barrières [Abortion care has too many barriers]. Medisch contact. 2019.

6. (NHG) NHG. NHG-Standpunt Effectiviteit en veiligheid van medicamenteuze overtijdbehandeling in de huisartsenpraktijk [NHG-Standard Effectivity and safety of medical abortion in general practices]. 2016.

7. (WHO) WHO. Safe abortion: technical and policy guidance for health systems. World Health Organization; 2012.

8. (NVOG) NVvOeG. Miskraam [Miscarriage]. 2020.

9. Chavkin W, Stifani BM, Bridgman-Packer D, al. e. Implementing and expanding safe abortion care: An international comparative case study of six countries. Int J Gynaecol Obstet. 2018;143 Suppl 4:3–11.

10. O’Connor R, O’Doherty J, O’Mahony M, Spain E. Knowledge and attitudes of Irish GPs towards abortion following its legalisation: a cross-sectional study. BJGP Open. 2019;3(4):bjgpopen19X101669.

11. Gaudu S, Crost M, Esterle L. Results of a 4-year study on 15,447 medical abortions provided by privately practicing general practitioners and gynecologists in France. Contraception. 2013;87(1):45–50.

12. Ho PC. Women’s perceptions on medical abortion. Contraception. 2006;74(1):11–5.

13. Newton D, Bayly C, McNamee K, al. e. ‘…a one stop shop in their own community’: Medical abortion and the role of general practice. Aust N Z J Obstet Gynaecol. 2016;56(6):648–54.

14. Godfrey EM, Thayer EK, Fiastro AE, al. e. Family medicine provision of online medication abortion in three US states during COVID-19. Contraception. 2021;104(1):54–60.

15. (IGJ) IgeJ. Jaarrapportage 2018 van de Wet afbreking zwangerschap [Year report 2018 of the Law termination of pregnancy]. Utrecht: Inspectie Gezondheidszorg en Jeugd; 2019.

16. Ferguson RM, Vanwesenbeeck I, Knijn T. A matter of facts… and more: an exploratory analysis of the content of sexuality education in The Netherlands. Sex Education. 2008;8(1):93–106.

17. Leusink P, Folkeringa-de Wijs M. De rol van de huisarts bij onbedoelde zwangerschap. Huisarts en wetenschap. 2017;60(6):298–301.

18. Goenee MS, Donker GA, Picavet C, Wijsen C. Decision-making concerning unwanted pregnancy in general practice. Fam Pract. 2014;31(5):564–70.

19. Beaman J, Schillinger D. Responding to Evolving Abortion Regulations — The Critical Role of Primary Care. New England Journal of Medicine. 2019;380(18):e30.

20. Michie S, van Stralen MM, West R. The behaviour change wheel: a new method for characterising and designing behaviour change interventions. Implement Sci. 2011;6:42.

21. Braun V, Clarke V. Using thematic analysis in psychology. Qualitative Research in Psychology. 2006;3(2):77–101.

22. Edwards PJ, Roberts I, Clarke MJ, Diguiseppi C, Wentz R, Kwan I, et al. Methods to increase response to postal and electronic questionnaires. Cochrane Database Syst Rev. 2009(3):Mr000008.

23. Batenburg R, van der Velden L, Vis E, Kenens R. Cijfers uit de registratie van huisartsen – een update van de werkzaamheidscijfers voor 2018 en 2019. Utrecht: Nivel; 2019.

24. Fiala C, Gemzel-Danielsson K. Review of medical abortion using mifepristone in combination with a prostaglandin analogue. Contraception. 2006;74(1):66–86.

25. Subasinghe AK, Deb S, Mazza D. Primary care providers’ knowledge, attitudes and practices of medical abortion: a systematic review. BMJ Sex Reprod Health. 2019.

26. Ganatra B. Health worker roles in safe abortion care and post-abortion contraception. Lancet Glob Health. 2015;3(9):e512–3.

27. Espey E, Ogburn T, Chavez A, al. e. Abortion education in medical schools: a national survey. Am J Obstet Gynecol. 2005;192(2):640–3.

28. Jackson CB. Expanding the pool of abortion providers: nurse-midwives, nurse practitioners, and physician assistants. Womens Health Issues. 2011;21(3 Suppl):S42–3.

29. Doran F, Nancarrow S. Barriers and facilitators of access to first-trimester abortion services for women in the developed world: a systematic review. J Fam Plann Reprod Health Care. 2015;41(3):170–80.

30. Sjöström S, Kopp Kallner H, Simeonova E, al. e. Medical Abortion Provided by Nurse-Midwives or Physicians in a High Resource Setting: A Cost-Effectiveness Analysis. PLOS ONE. 2016;11(6):e0158645.

31. Lipp A. Stigma in abortion care: application to a grounded theory study. Contemp Nurse. 2011;37(2):115–23.

32. de Moel-Mandel C, Shelley JM. The legal and non-legal barriers to abortion access in Australia: a review of the evidence. Eur J Contracept Reprod Health Care. 2017;22(2):114–22.

33. Schonberg D, Wang LF, Bennett AH, al. e. The accuracy of using last menstrual period to determine gestational age for first trimester medication abortion: a systematic review. Contraception. 2014;90(5):480–7.

34. Kaneshiro B, Edelman A, Sneeringer RK, Ponce de Leon RG. Expanding medical abortion: can medical abortion be effectively provided without the routine use of ultrasound? Contraception. 2011;83(3):194–201.

