## Supplementary material for "Are Dutch General Practitioners willing to prescribe mifepristone and misoprostol?: a mixed-methods study": Decision of the Research Ethics Review Committee VU Amsterdam

We thank you for your time spent taking this survey.
Your response has been recorded.

Below is a summary of your responses

Download PDF

Welcome to the ethics review self-check provided by the FSW Research Ethics Review Committee (RERC).

In this self-check you will be asked a series of questions to test whether your research complies to the guidelines in the FSW's research code of conduct.

If your research is "standard" and conforms to these guidelines, you will receive a confirmation message and further review by the RERC is not required.

In "standard" research:

- No harm is envisaged for the participants or the population from which participants have been drawn
- Participants receive complete and accurate information about the goals of the research before they participate
- Participants give active consent for participation in the research
- Participants are not deceived without being thoroughly debriefed
- Participants are healthy adults who are not in a vulnerable position
- Personal and sensitive data are kept confidential and are stored in a secure environment.

If your research deviates from these standards, further ethics review may be required. Proceed to the self-check to determine whether this is the case. If you already know your research is non-standard, you may proceed to the full ethics review form below.

If your research is of a medical nature, poses a medical risk, or includes invasive procedures, ethics review is **always** required. In this case, your research should be evaluated by the medical-ethical review committee (METC).

evaluated by the medical-ethical review committee (METC).

● **Proceed with the self-check (and click ">>" below)**

---

*Other forms:*

[Proceed directly to the full ethics review form]

*Only choose this option if you are sure your research is not "standard"!*

[Proceed to the medical ethics review committee]

*Only choose this option if you are sure your research is medical in nature!*

---

Your name:

| | |
|---|---|
| First name | Julia |
| Family name | Schellekens |

---

Your email address:

---

Your VU net ID:

jss384

---

Your position:

Student in a master program

---

Name of (primary) supervisor:

Rebecca Gomperts

---

Department:

Other

---

Other department:

[ ]

Name of your research project:

Willingness of the general practitioner to provide the abortion pill

Is an ethics review for the present research project required by a**third party**? (*)
*A third party may be a funder (e.g., NWO, ERC), a journal, or another party outside VU Amsterdam.*

- ○ No
- ● **Yes**

*(*) This question helps us to determine whether you will need a written statement by the Research Ethics Review Committee, or whether an automated e-mail suffices.*

Will you collect or analyze **data on individuals** in the course of your research project?
*Data include interview and survey responses, observations, registrations, social media content, and visual images.*

- ○ No (this will end the self-check; no ethics review is necessary)
- ● **Yes**

Is the data you use collected in the public or in the private domain?
*Public domain data includes a.o.: observations made during a public event, observations of public figures, or data extracted from public online sources (including publicly accessible social media data).*
*Private domain data includes a.o.: observations made using (online) questionnaires, experiments, and interviews, as well as data extracted from private digital sources.*

- ○ Public domain
- ● **Private domain**
- ○ Both

Do the data you use contain personal or sensitive information?

*Personal data contains information that may identify a specific person, e.g. name, address, phone number, IP address, bank account number, social security number (BSN).*
*Sensitive data contains information such as race, religion, sexual orientation, criminal record, and political preference.*

- ○ No
- ● **Yes (*)**

---

*(*) Note that personal or sensitive data should be protected and not be distributed to others.  Data should be stored at a secure location (i.e., your home-drive at the VU; not a portable hard drive or USB stick). Access should be limited by use of encryption software (i.e. Bitlocker, 7-Zip). Also, particular attention should be given to the design of your research data management plan.*

---

Will the research project involve merging multiple data sets with information on individuals?
Note: merging multiple waves of the same longitudinal study is not considered merging multiple data sets.

- ● **No**
- ○ Yes (*)

---

*(*) Merging multiple data sets may pose non-anticipated risks to participants. Therefore the RERC advises a full ethics review in case data sets are merged.*

---

Will you ask participants in the research project for **informed consent**?

*Note that in the case of minors (participants below the age of 16), informed consent should be obtained from a parent or legal caregiver, as well as from the minor. In the case of participants between the ages of 16 and 18, consent should be obtained from the participant, and the parents/legal caregivers should be informed.*

- ○ No (*)
- ● **Yes**

---

*(*) Full ethics review will be required to determine whether in your study informed consent procedures may be skipped*

---

*The questions below are about potential risks envisioned for your research participants. Note that the term "participant" refers to any human subject about whom observations are being made, regardless of whether this happens in the public or private domain.*

Does the research you plan to conduct pose potential risks to the **participants** during or after the research?
*Risks may include physical and psychological harm or discomfort.*

- ● **No**
- ○ Yes (*)

Does the research you plan to conduct pose potential risks to a **population or group** from which your participants are drawn?
*Risks may include stigmatization, or reputational or economical damage.*

- ● **No**
- ○ Yes (*)

*(*) Further ethics review will be required.*

Are your participants individuals who are **vulnerable**?
*Participants may be vulnerable when they depend on others for assistance in daily life, or when they experience threats or physical danger.*

- ● **No**
- ○ Yes (*)

*(*) Further ethics review will be required.*

Will participants be exposed to material, social, or psychological **recruitment incentives** that are stronger than usual?
*E.g., are payments made that are higher than the minimum wage? Are individuals exposed to strong social or psychological pressure to participate?*

- **No**
- Yes (*)

---

*(*) Further ethics review will be required.*

---

Will participants be exposed to **research stimuli** (e.g., pictures, video, text) that may be distressing, offensive, or age-inappropriate?
*For minors, age guidelines for media content (e.g., de Kijkwijzer) apply. For participants over 18, research stimuli may be considered distressing or offensive if they are stronger than what participants would normally be exposed to in daily life.*

- **No**
- Yes (*)

---

*(*) Further ethics review will be required.*

---

Does the research you plan to conduct pose potential risks to the **researchers**?
*Risks may include physical and psychological harm or discomfort.*

- **No**
- Yes (*)

---

---

Will you **deceive** participants in the research?
*Deception means that you deliberately give participants information that is false.*

- **No / Yes, but they will be thoroughly debriefed afterwards**
- Yes, and they will not be debriefed (*)

---

*(*) Further ethics review will be required*

---

Do you guarantee **anonymity or confidentiality** to your participants?
*Anonymity means that the identity of participants will remain unknown, also to you. Confidentiality means that you know the identity of participants, but it will be unknown to others.*

- ○ No (*)
- ○ Yes, participants will be anonymous
- ● **Yes, participant data will be kept confidential (**)**

---

*(*) Further ethics review will be required*

*(**) Note that in this case particular attention should be given to the design of your research data management plan.*

---

Will you conduct your research, or collect data, outside of the Netherlands ?

- ● **No**
- ○ Yes (*)

---

*(*) Note that in this case you should make sure that you are aware of, and comply to, local guidelines for research ethics.*

---

### Result of the research ethics self-check:

---

Your research project **does not require further evaluation** by the Research Ethics Review Committee. This is because:

- You have indicated that you will ask your participants for informed consent.
- You have indicated that your research poses no risks to participants.
- You have indicated that you will not work with participants who are vulnerable.
- You have indicated that your participants are not exposed to material, social or psychological recruitment incentives that are stronger than usual.
- You indicated that your participants will not be exposed to research material that is distressing, offensive, or age-inappropriate.
- You have indicated that your research poses no risks to the researchers.
- You have indicated that you will not deceive research participants, or you will properly debrief them afterwards.
- You have indicated that respondents in your research will be fully anonymous.

What to do now?
If you think these results are mistaken, please go back in the form and change your entries accordingly.

If the results are correct, we ask you to confirm this, after which they will be stored in our repository. You will also get a receipt, displaying all answers you provided, for future reference.

---

Additional note: Research data management plan required

- You have indicated that you collect personal / sensitive data, and/or that the identities of the participants are known to you. This means that you will need to pay particular attention to the research data management plan (DMP) for your research. For more information on how to draft your DMP, please visit the Faculty's page on VUNet about data management contact the Faculty's Data Steward, Koen Leuveld.

---

You have completed the ethics review self-check.

● **I have completed this form truthfully.**

---

Click below to receive a confirmation message on the following screen, displaying your responses. Click on the pdf icon in the top right corner to save your confirmation for future reference.

○ Proceed (and click ">>" below)

Powered by Qualtrics
